# Quantitative biomarker profiling of serum samples in the By-Band-Sleeve trial

**DOI:** 10.1101/2025.11.11.25339993

**Authors:** Madeleine L. Smith, Lucy J. Goudswaard, David A. Hughes, Jane M. Blazeby, Chris A. Rogers, Graziella Mazza, Eleanor A. Gidman, Sophie Fitzgibbon, Alix Groom, Susan M. Ring, Nicholas J. Timpson, Laura J. Corbin

## Abstract

Metabolomics data has been generated via proton nuclear magnetic resonance (NMR) spectroscopy in samples collected within By-Band-Sleeve. Two sample collection efforts were made – firstly, from a randomised controlled trial (RCT) comparing the effectiveness of three types of bariatric surgery: the Roux-en-Y gastric bypass (“bypass”), laparoscopic adjustable gastric band (“band”) and the sleeve gastrectomy (“sleeve”), and secondly from a non-randomised (observational) study of bariatric surgery. In both instances, samples were collected from patients before and after surgery. Data underwent quality control (QC) using a standard pipeline via the R package *metaboprep*. This package extracts data from preformed worksheets, provides summary statistics and enables the user to select samples and metabolites for their analysis based on a set of quality metrics. Post-filtering, the dataset consists of data from 1410 samples (999 pre-surgery, 411 post-surgery) from 1045 unique individuals (1000 from the RCT and 45 from the non-randomised study), each with 250 measured metabolic traits. Comparison of NMR measures to clinical chemistry data showed good agreement for the metabolites in common across both datasets. Concordance with previous NMR data generated for a subset of the same samples was largely good. Overall, this data note describes the data, explains the pre-processing and quality control procedures applied to the data, and provides some data validation analyses.

## Introduction

By-Band-Sleeve (BBS) is a UK-based randomised controlled trial (RCT) comparing the clinical and cost-effectiveness of three types of bariatric surgery - Roux-en-Y gastric bypass (“bypass”), adjustable gastric banding (“band”), and sleeve gastrectomy (“sleeve”)^1,2^.

Quantifying metabolites and small molecules (known as metabolomics) in samples collected within RCTs of weight loss interventions allows exploration of the biological response to weight change, which is known to broadly perturb cellular metabolism^3,4^. Furthermore, the randomised design of BBS can be harnessed to compare the metabolomic effects of gastric bypass, gastric band and sleeve gastrectomy, providing molecular insight into their differential modes of action. This data note describes the metabolomics data collected in BBS via proton nuclear magnetic resonance (NMR) spectroscopy.

The NMR metabolomics data in BBS were generated by Nightingale Health. This high-throughput platform simultaneously quantifies the cholesterol and triglyceride content of lipoprotein subclasses, as well as their size and concentration; amino acids (aromatic and branched-chain); fatty acids; glucose and pre-glycaemic factors (including lactate and citrate); fluid balance molecules (including albumin and creatinine); glycoprotein acetyls (a marker of inflammation); and other lipids (including cholines and sphingomyelins). Although it measures a smaller set of metabolites compared to some mass spectrometry platforms, one advantage of this NMR platform is that the metabolites are quantified in absolute concentration units, providing physiologically and clinically meaningful data. This enables better exploration of the relationship between the measured metabolites and an individual’s health status^5^. Further, the absolute nature of the data means that metabolite levels can be compared across studies.

This data note describes the NMR data collected within BBS and explains the quality control (QC) and pre-analytical processing applied to the data, including some validation analyses. The mass spectrometry metabolomics data generated within BBS has previously been described^6^.

## Materials and methods

### Overview of data sources By-Band-Sleeve

BBS is an open parallel-group multi-site randomised controlled trial which took place in the UK (registration number: ISRCTN00786323)^1,2^. The trial investigated the clinical and cost-effectiveness of three types of bariatric surgery: the Roux-en-Y gastric bypass (“Bypass”), laparoscopic adjustable gastric band (“Band”) and the sleeve gastrectomy (“Sleeve”). This study began in December 2012 as “By-Band” comparing Band and Bypass, the two most common bariatric surgery methods in the UK at that time. However, due to the increasing number of sleeve gastrectomy procedures taking place in the UK during the first few years of the study, Sleeve was added to the study which became “By-Band-Sleeve” in August 2015^7^. The pilot phase of the study occurred in two study sites (designated below as A and B), expanding into ten more sites (labelled C-L) after the adaptation to include sleeve. Ten of the twelve sites agreed to collect and store biological samples for future research (see below). At completion of recruitment in September 2019, 1351 patients had been randomised as part of the trial^8^. The primary endpoint for the trial was 36-months post-randomization and follow-up to this timepoint was completed by the end of March 2023 with the main trial outcome reported in May 2025^1^.

### By-Band-Sleeve non-randomised metabolomics sub-study

When recruitment to the main BBS trial concluded (September 2019), recruitment to a non-randomised metabolomics sub-study began and ran until June 2022, at which point 214 patients had been consented. Eligibility criteria remained the same as in the main trial, but participants of the sub-study were not randomised to a surgery type and instead underwent the surgical procedure as determined by the usual process at their hospital with follow-up care as standard. Participants were followed up for 12 months after their operation. Seven of the original 12 study sites opted to recruit patients for the sub-study, including site L which had not collected samples for the main trial. A reduced set of clinical data were collected for the sub-study participants, using a different data collection form to the main study that focused on weight and diabetes outcomes pre-surgery and at follow-up. Herein, the two parts of BBS described will be referred to as the main trial and the sub-study.

### By-Band-Sleeve – derived variables

Variables were derived to capture the differences in timing of surgery and sample collection across participants. Time between baseline sample collection and surgery date, time between surgery date and follow-up sample collection, and time between baseline and follow-up sample collection were calculated in days and presented to the nearest month. Storage time was calculated as the difference between the date the sample was collected and the date it was thawed for aliquoting prior to metabolomics analysis, in days to the nearest month. Disruption by the COVID-19 pandemic meant that adherence to the BBS trial protocol (especially with respect to the format and timing of the follow-up appointments) was interrupted in some cases. For example, some of the 36-month appointments were conducted over the phone, with bloods collected at a later date when COVID-19 restrictions would allow.

### Ethical approval

This study was conducted in accordance with the principles of the Declaration of Helsinki.

Ethical approval for BBS was granted by the Southwest Frenchay Research Ethics Committee (reference 11/SW/0248) and written informed consent was obtained from all participants. All samples were used, stored and disposed of in accordance with the Human Tissue Act 2004.

### Sample collection

During the BBS main trial, data collection points were scheduled at baseline (pre-randomisation), the day of surgery, four weeks post-surgery, and 6-, 12-, 24- and 36-months post-randomisation. Separately to the work presented here, blood samples were taken at each timepoint (excluding the day of surgery) for hospital laboratory analyses including, for example, full blood count, liver function tests and HbA1c (haemoglobin A1c, measure of glycated haemoglobin) measurement. Patients were asked to fast for a minimum of four hours prior to blood collection. In addition, blood samples were collected at baseline and 36-months post-randomisation and used to isolate whole blood, plasma and serum samples for future research. From this collection, serum samples were used to generate metabolomics data. Blood was collected into 4ml vacutainers containing clot activator gel (Serum Separation Tubes (SST)) and processed within one hour of collection when possible. The blood was allowed to clot for 30 minutes before centrifugation at 1500g for 10 minutes at room temperature to separate the serum. The serum was transferred into sterile polypropylene tubes with screw cap using a Pasteur pipette or similar and stored at -80°C before being shipped to the coordinating centre (University of Bristol) on dry ice.

In the BBS non-randomised sub-study, fasting blood samples were collected pre-surgery and 12 months post-surgery and used to isolate serum, following the same protocol as the main trial. The serum was transferred into four sterile polypropylene tubes with screw cap with a target sample volume of 0.5ml per tube. These blood samples were stored at -80°C before being transferred from each recruiting site to the University of Bristol for storage.

From the main trial, 1651 samples from 1149 participants collected between January 2013 and June 2022 were sent for NMR metabolomics analysis by Nightingale Health, Finland. These were all the samples available in Bristol at the point of shipping to Nightingale; not all participants had both a baseline and a 36-month sample. From the sub-study, 45 pairs of samples (90 samples; baseline and 12-month samples) collected between October 2019 and May 2022 were sent for NMR metabolomics analysis; these were all the pairs of samples available in Bristol at the point of shipping. The total number of BBS samples (main trial and sub-study) analysed by Nightingale was 1741. Each study site had different numbers of samples available at the point of shipping, and therefore the number of baseline, endpoint and paired samples differs across the sites (**Table 1**).

**Table 1.**
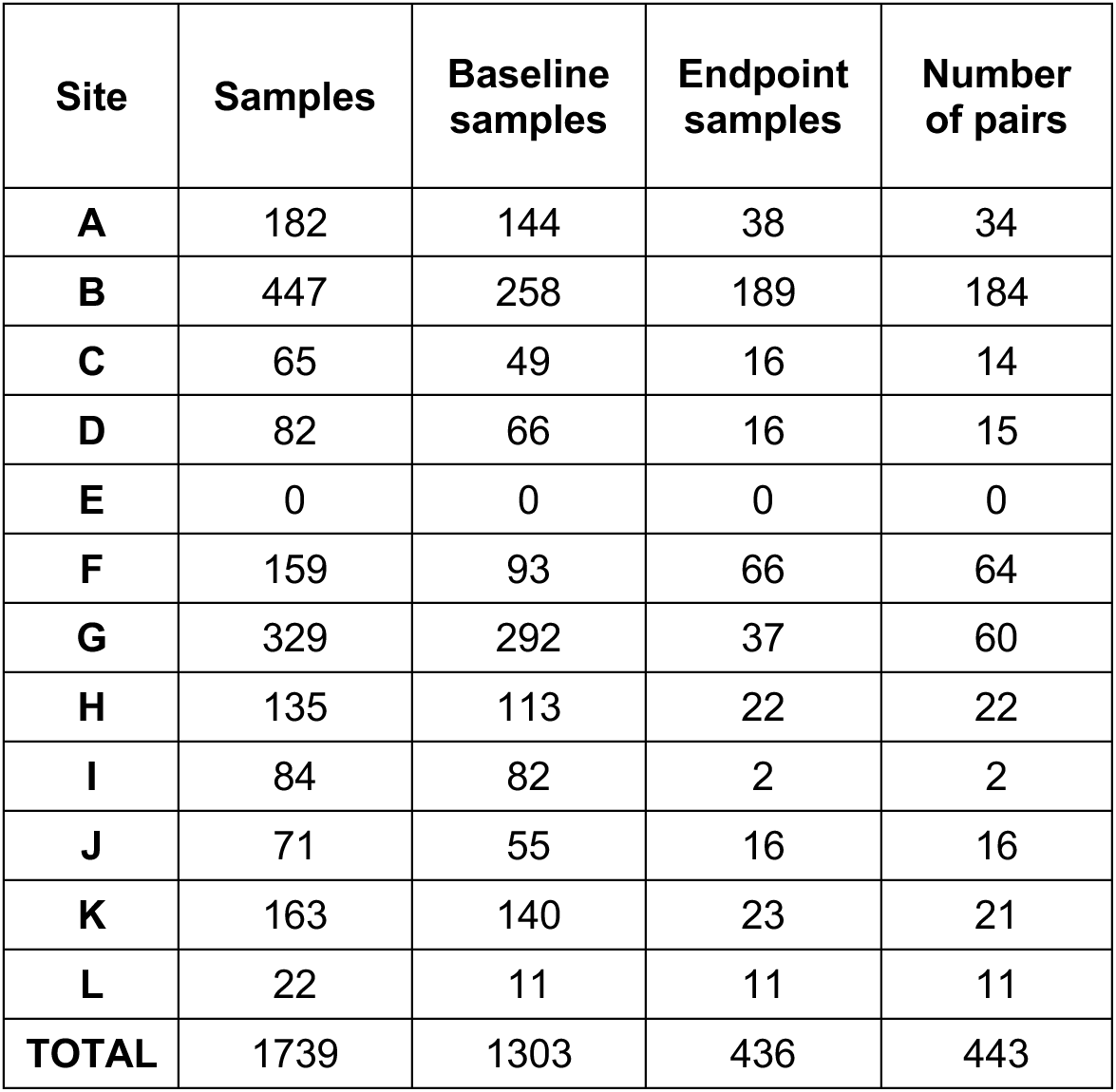
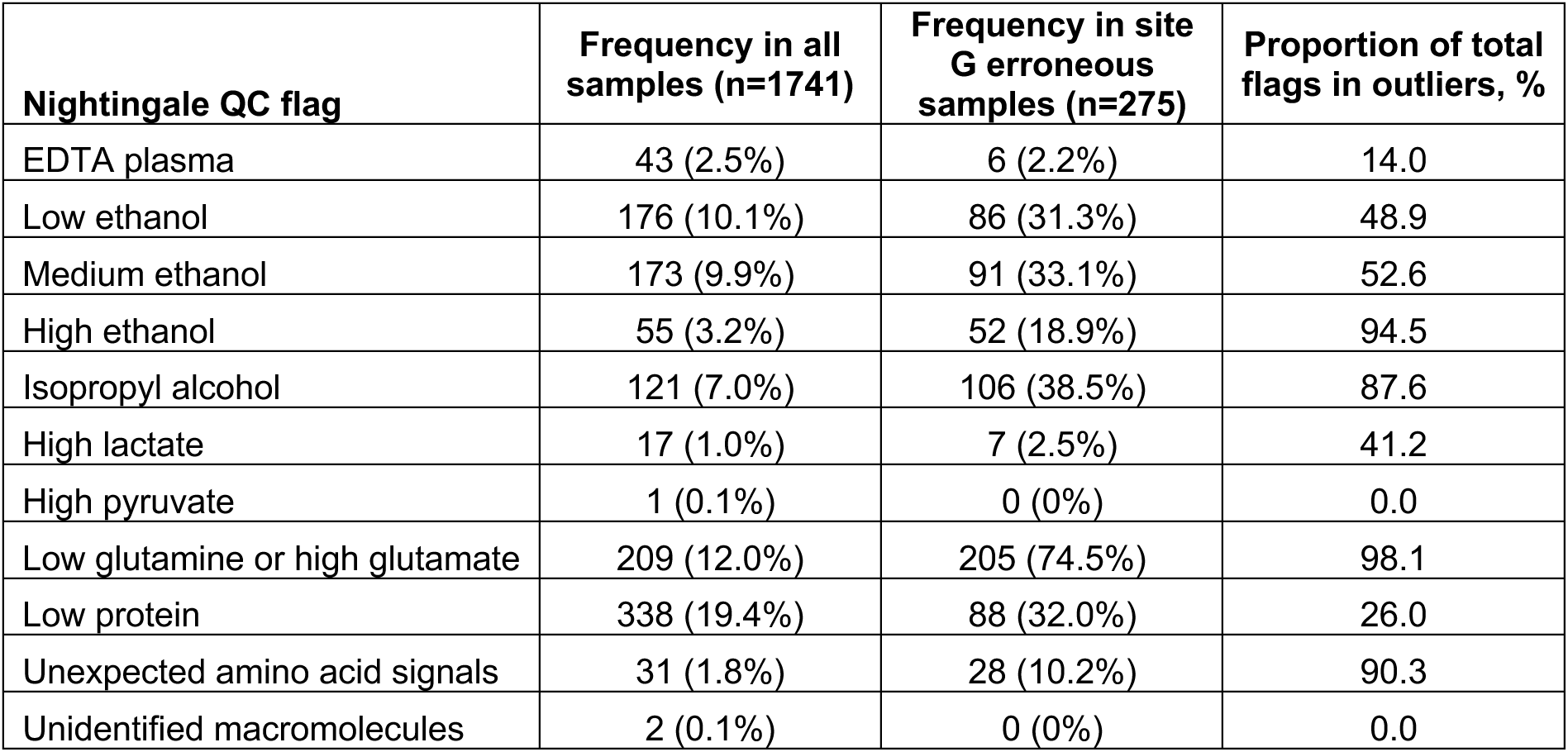
The distribution of BBS samples across the 12 study sites (main trial and non-randomised sub-study).

### Metabolite data acquisition

A high-throughput ^1^H-NMR metabolomics platform was used to quantify 250 metabolic biomarkers, 170 in absolute levels and 80 ratio measures. The biomarkers include detailed measures of cholesterol metabolism, fatty acid compositions, and various low-molecular weight metabolites, such as amino acids, ketones and glycolysis metabolites. Details of experimental procedures have been described elsewhere^9,10^. Data were returned in July 2023.

### Pre-analytical processing

#### Duplication of previously identified sample outliers

Aliquots of samples analysed by NMR had been previously analysed by Metabolon using mass spectrometry. During the pre-processing of the Metabolon data, a group of outlying and potentially erroneous samples were identified by principal component analysis (PCA), all from site G^6^. With this in mind, when preparing the samples for NMR, duplicates of all the samples that had previously been identified as outliers on the Metabolon platform were also sent for analysis. This would allow us to know if the error was specific to the aliquot analysed for that participant previously or was a property of the parent sample (and would therefore affect all aliquots derived from that same sample). If the problem was with the individual aliquots, then the duplicate samples would show an improvement in quality.

We compared these duplicate samples using Bland-Altman plot analysis ^11,12^ (R package: *blandr*^13^ with default settings) to assess concordance. We used PCA and Student’s t tests to assess whether the NMR data also showed that these samples were outliers. PCA was performed using a subset of independent features. Independent features were identified by constructing a dendrogram based on a Spearman’s rho distance matrix, where distance is calculated using the absolute value of 1-Spearman’s rho. A set of k-clusters were identified using tree cut height of 0.5. Within each k-cluster, one representative feature was chosen to represent the cluster. It was identified as the feature with least missingness or chosen at random in the case of missingness ties. Only for the purpose of performing the PCA, missing values in this set of independent features were imputed to the median before the data were standardised (z-transformed) such that the mean was equal to zero and the standard deviation was equal to one for each feature. Student’s t tests were performed to compare the levels of each metabolite in the group of potentially outlying samples to the rest of the samples. The same test was carried out using 1000 random subsets containing the same number of samples and the median number of metabolites with differences in mean across the 1000 iterations calculated.

#### Quality control and filtering using *metaboprep*

The R package *metaboprep*^14^ was used to perform pre-processing and quality control of the data, after excluding the previously identified outliers and their duplicates (described above). Both sample missingness and feature missingness were considered when filtering the data. Any samples and features with extreme missingness (>80%) were excluded before recalculating sample and feature missingness. Then samples with more than 20% missingness and features with more than 50% missingness were removed from the dataset. PCA analysis was also performed by *metaboprep* (as described above) and any samples that were more than five SDs from the mean of the first and/or second principal components were filtered from the data set.

### Data validation

#### Comparison to clinical chemistry

We explored concordance between NMR-derived measures and clinical chemistry measures for the metabolites that were assessed by both methods. The clinical chemistry assays were performed by the hospitals (using standard procedures) on samples collected at the same time as the samples that were frozen and later sent for NMR analysis. The metabolites that were compared are: creatinine, glucose, albumin, total cholesterol, LDL cholesterol, HDL cholesterol and total triglycerides. BBS sub-study samples did not undergo clinical chemistry assays, so the comparisons were done for main trial samples only. We compared the NMR data to clinical chemistry using Bland-Altman plot analysis^11–13^. One outlier was removed from the clinical fasting glucose variable before plotting as it was assumed to be a reporting error (76 mmol/L).

#### Comparison to previously generated NMR data

A subset of 250 samples from 125 site B participants in the BBS main trial were analysed using the same NMR platform in 2022 as a pilot experiment (data returned in October 2022). In this instance, samples were analysed by the NMR facility at University of Bristol and raw data sent for quantification by Nightingale Health, Finland. These 250 samples were also included in the current sample set and have therefore been analysed twice. When sending the samples for the second time, different aliquots from the same sampling occasion were used, to ensure that they had undergone the same number of freeze/thaw cycles. We compared the data from each run (2022 vs. 2023) using Bland-Altman plot analysis^11–13^.

## Results

### Participant characteristics

The baseline characteristics of all BBS main trial participants have been published^8^. The characteristics of the subsample of 1192 main trial participants and 45 sub-study participants whose samples were analysed by Nightingale are described in **Table 2**.

**Table 2.** Participant characteristics for the BBS main trial and sub-study.

| Characteristic | N<sup>1</sup> | Overall, N = 1,192<sup>2</sup> | Main, N = 1,147<sup>2</sup> | Sub-study, N = 45<sup>2</sup> |
| --- | --- | --- | --- | --- |
| Sex | 1,167 |  |  |  |
| Male |  | 285 (24%) | 279 (25%) | 6 (13%) |
| Female |  | 882 (76%) | 843 (75%) | 39 (87%) |
| Age (years) | 1,166 | 47 (11) | 47 (11) | 45 (10) |
| Ethnicity | 1,167 |  |  |  |
| White |  | 1,008 (86%) | 963 (86%) | 45 (100%) |
| Mixed/multiple ethnic groups |  | 25 (2.1%) | 25 (2.2%) | <5 |
| Asian/Asian British |  | 58 (5.0%) | 58 (5.2%) | <5 |
| Black/African/Caribbean/Black British |  | 65 (5.6%) | 65 (5.8%) | <5 |
| Other |  | 11 (0.9%) | 11 (1.0%) | <5 |
| Diabetes status | 1,167 |  |  |  |
| No indication |  | 811 (69%) | 773 (69%) | 38 (84%) |
| Glucose impairment |  | 13 (1.1%) | 13 (1.2%) | <5 |
| Oral hypoglycaemics |  | 227 (19%) | 223 (20%) | <5 |
| Insulin |  | 56 (4.8%) | 56 (5.0%) | <5 |
| GLP-1 agonist |  | 25 (2.1%) | 23 (2.0%) | <5 |
| Diet controlled |  | 35 (3.0%) | 34 (3.0%) | <5 |
| Baseline<sup>3</sup> weight (kg) | 1,167 | 130 (23) | 130 (24) | 126 (21) |
| End<sup>4</sup> weight (kg) | 1,011 | 103 (26) | 103 (26) | 93 (22) |
| Baseline<sup>3</sup> BMI (kg/m<sup>2</sup>) | 1,167 | 47 (7) | 47 (7) | 46 (6) |
| End<sup>4</sup> BMI (kg/m<sup>2</sup>) | 1,011 | 37 (8) | 37 (8) | 34 (6) |
| Diabetes duration (months)<sup>5</sup> | 313 | 74 (70) | 74 (71) | 43 (34) |

<sup>1</sup>N with data.
<sup>2</sup>For continuous variables values are mean (SD), for categorical variables values are n (%). <sup>3</sup>'Baseline' refers to the first pre-surgery measures; these were taken at randomization in the main trial and at the first visit in the non-randomised sub-study. <sup>4</sup>'End' refers to the measures taken at 36-months post-randomization in the main trial and at 12-months post-surgery in the non-randomised sub-study.
<sup>5</sup>Diabetes duration at the time of recruitment
<5 may include zero

### Observations relating to sample quality

An overview of results (**Supplementary File 1**) and quality report (**Supplementary File 2**) were provided by Nightingale Health. A total of 72 samples had no data returned (i.e., all NA); these samples had QC flags of ‘technical error’ (n=15), ‘solid material’ (n=8), ‘missing samples’ (n=3) or ‘diluted’ (n=46). Forty-three samples were identified as containing ethylenediaminetetraacetic acid (EDTA), which could mean that they were plasma samples rather than serum. Nightingale recommends that the pyruvate, glycine and glycerol measures for these samples are disregarded, as these will be affected by the presence of EDTA. Other contaminants detected in samples included ethanol and isopropyl alcohol. Ethanol can distort glycerol, 3-hydroxybutyrate, acetate and valine concentrations whilst isopropyl alcohol can distort creatinine concentrations. Therefore, caution is recommended when interpreting the concentrations of these metabolites for the contaminated samples. Some of the samples have been flagged as having low glutamine or high glutamate, which is a common sign of sample degradation. In these samples glutamine is not quantified, but this may also interfere with pyruvate quantification. 338 samples were found to have low protein, which indicates potential sample dilution, and therefore caution is recommended when analysing absolute concentrations from these samples.

### Pre-analytical processing

**Figure 1** provides an overview of the of pre-analytical data processing and filtering applied to the data set. The (n=142) samples previously identified as outliers - based on data produced from the Metabolon platform and previously published^6^ - and that were, here, analysed in duplicate (see Methods) showed good agreement within duplicate pairs (**Figure S1 in Supplementary File 3**). This suggests that the technical artefact causing them to be different from the rest of the sample population was not specific to one aliquot but a property of the parent sample. PCA showed that the previously identified outlying samples and their duplicates clustered together but still overlapped with the main cluster of samples (**Figure 2**). After Bonferroni correction for multiple testing, 206 out of 250 (82.4%) metabolites showed a difference in mean concentration between the potentially outlying samples from site G and all other samples in the dataset (p<0.05/250, **Table S1 in Supplementary File 4**). Across 1000 randomly selected groups of 140 samples (to match the number of outliers after removal of two samples with no data returned), the median number of metabolites with differences in mean (p<0.05/250) between the selected group and the rest of the samples was 0 (range 0-9).

**Figure 1.**
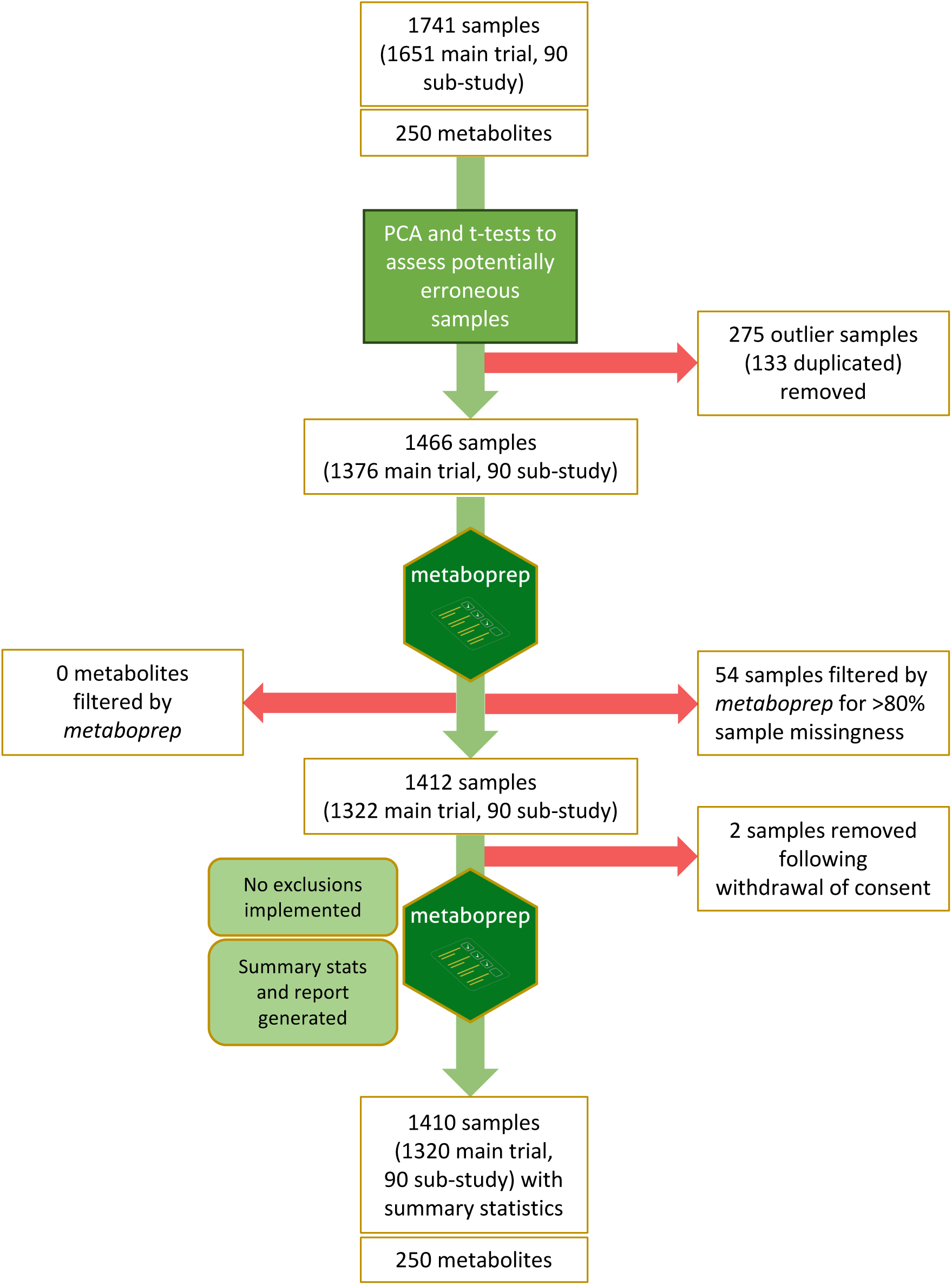
Overview of pre-analytical processing and data filtering. The R package *metaboprep* pipeline was implemented to filter the samples and metabolites based on missingness, principal components and total peak area. Finally, the pipeline was run again on the final datasets separately but without exclusions applied, to generate summary statistics for the final data set.

**Figure 2.**
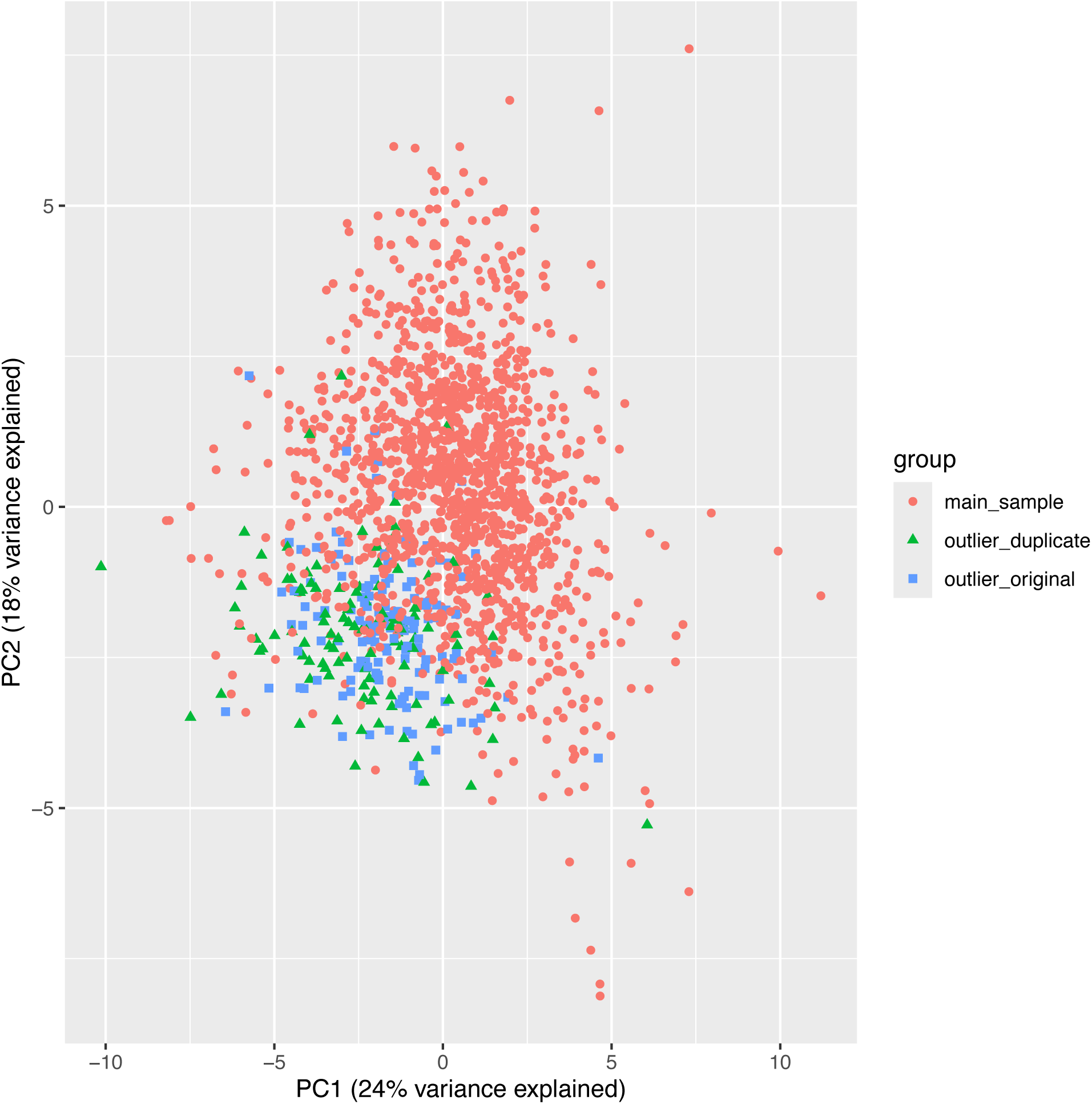
Principal component (PC) plot of PCs 1 and 2 (calculated in the NMR data) with samples coloured by whether they were an outlier (as identified in the Metabolon data) (*outlier_original*), a duplicate of an outlier sample (*outlier_duplicate*) or neither (*main_sample*).

Many of the samples with QC flags (see previous section) were among the outliers from site G. **Table 3** summarises the sample quality flags in the full data set (1741 samples) and the outlying samples from site G (275 samples). More than 85% of the samples with the flags ‘high ethanol’, ‘isopropyl alcohol’, ‘low glutamine’ or ‘high glutamate’, and ‘unexpected amino acids’, were in the site G outlier group. Based on these observations, we believe these samples may be outliers because of a technical artefact, e.g., sample processing differences, and recommend that researchers exclude these Site G samples from statistical analyses. Here we removed the 142 original outliers and their duplicates (total of 275 samples) from the data set before further filtering and QC were carried out.

The remaining 1466 samples were QC filtered using *metaboprep*^14^. Based on having extreme sample missingness (>80%) missingness, 54 samples were removed from the data set. These samples were QC fails recorded by Nightingale and set entirely to missing (NA) prior to the return of the data (see above). No samples or features were excluded for any other reasons within the *metaboprep* pipeline. Following the removal of two BBS participants who had subsequently withdrawn consent for their data to be used, the final filtered data set comprised 1410 samples (1320 main trial and 90 sub-study samples; 999 pre-surgery and 411 post-surgery) from 1045 unique individuals (1000 main trial / 45 non-randomised sub-study) and 250 metabolites (170 absolute measures and 80 ratios). The *metaboprep* summary report provides summary statistics for the final dataset (**Supplementary File 5**).

### Data validation

#### Comparison to clinical chemistry

The NMR-derived measures mostly showed good agreement with the clinical chemistry data, although concentrations tended to be lower in the NMR data and proportional bias was evident in some cases, e.g., albumin, total triglycerides (**Figure 3**). Removing samples with the ‘low protein’ flag, indicative of poor sample quality, led to better agreement across all metabolites (**Figure S2 in Supplementary File 6**).

**Figure 3.**
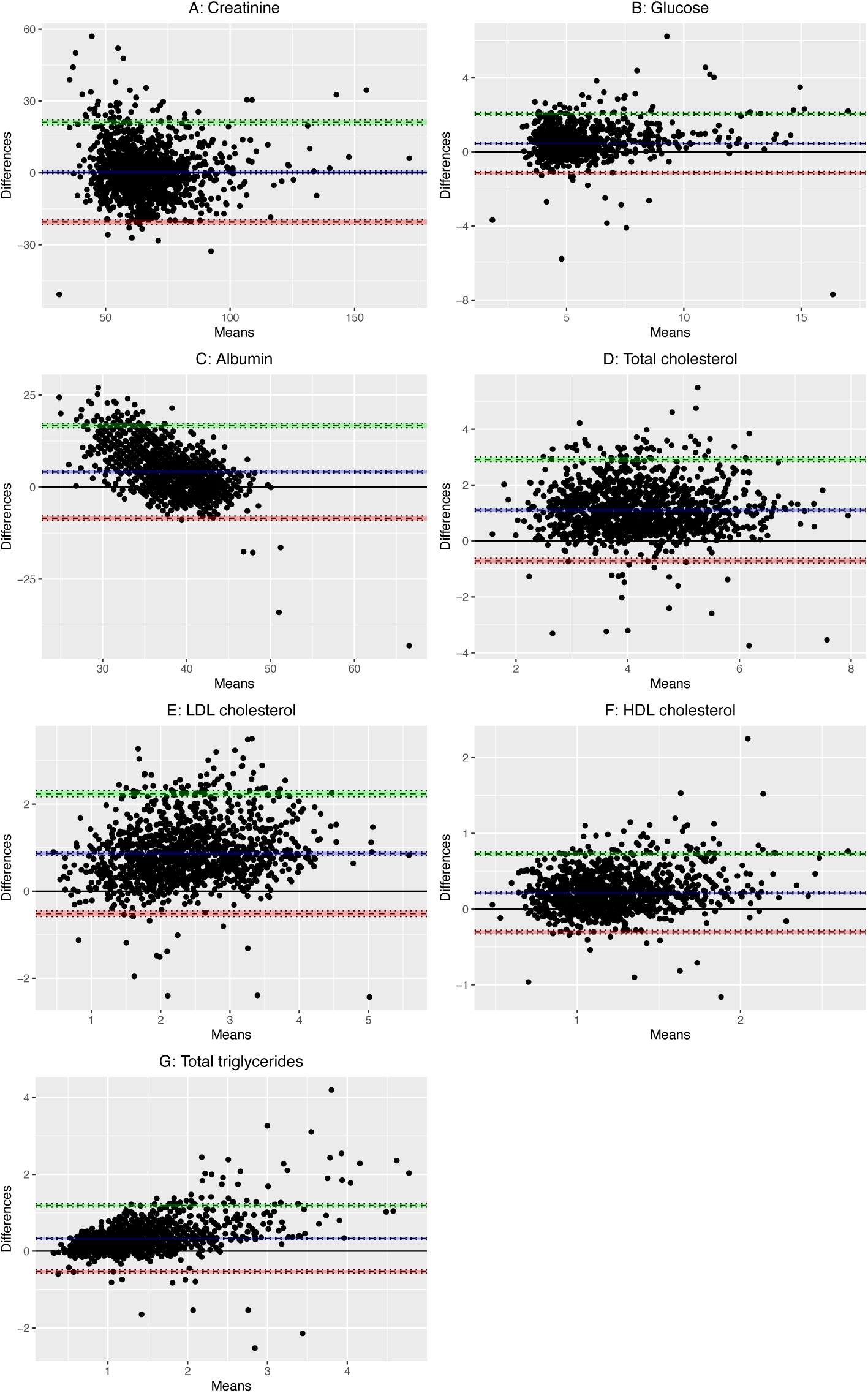
Bland-Altman plot comparison of (A) creatinine (n=1200), (B) glucose (n=1184), (C) albumin (n=1229), (D) total cholesterol (n=1228), (E) LDL cholesterol (n=1200), (F) HDL cholesterol (n=1224) and (G) total triglyceride (n=1212) concentrations measure by NMR versus clinical chemistry. LDL, low density lipoprotein; HDL, high density lipoprotein; NMR, nuclear magnetic resonance.

#### Comparison to previous BBS NMR data

The agreement between measures derived from the previous pilot study conducted using 125 pairs of samples from site B to the dataset described herein was generally good (**Figure S3 in Supplementary File 7**). Similarly to the comparison with clinical chemistry derived measures, there was evidence of proportional bias for some metabolites. Where there was evidence of bias, there was a tendency for values measured in the pilot to be greater than those measured as part of this larger analysis.

## Discussion

In this data note we have provided an overview of the NMR metabolite data generated within BBS, including details of the pre-analytical data processing, QC procedures, participant characteristics and data validation.

Samples that had been previously identified as outliers in the Metabolon data^6^, all from the same study site (site G) were not obvious outliers in the NMR data, as determined by visual inspection of the PCA. However, further investigation of the NMR data showed differences in this subset of samples that led us to exclude them from the dataset. While the structure of this error is not likely to be directly related to our primary analysis focus (looking for differences between pre- and post-surgery), the samples in this outlier group were almost all pre-surgery samples so they have the potential to induce bias. Based on the NMR data alone we would likely not have identified this group as being potentially problematic. This may be because the metabolites measured by mass spectrometry (over 1000 metabolites) were better able to capture the differences in sample handling procedures that we presume affected this group of samples. However, the sample quality flags provided alongside the NMR data provided an indication that these samples were of lower quality.

When comparing the metabolites in common across NMR and clinical chemistry assays there was good overall concordance that improved following removal of potentially poor-quality samples. However, for some metabolites there was evidence for proportional bias, including for albumin. This is in keeping with data in the UK Biobank, where the correlation coefficient between NMR- and clinical chemistry-derived albumin was weaker than observed for the other metabolites^15^. The fact that samples analysed by NMR had been stored in the freezer prior to analysis whereas clinical chemistry assays were typically run on fresh bloods may have contributed to the differences in concentration by the two methods. Albumin and a small number of other metabolites also showed some directional bias when comparing data from a subset of samples that had previously been analysed. The lack of consistency observed for these metabolites may be due (at least in part) to samples being run in different spectrometers. The differences in setup between the Bristol NMR lab (using 600MHz NMR for one spectrometer) and Nightingale’s internal NMR labs (all 500 MHz) are anticipated to enhance such deviations for duplicates. Given our findings, users may want to consider excluding samples from the current dataset based on specific sample quality flags, in particular the ‘low protein’ flag which can indicate that sample dilution has occurred.

The NMR metabolomic data in BBS provides the opportunity to explore the biological mechanisms that underpin how weight loss via bariatric surgery influences health. In addition, data from BBS can be compared to populations undergoing other weight loss interventions such as caloric restriction, which could allow differentiation between the metabolomic effects of weight-loss itself, and intervention-specific effects.

### Strengths and limitations

BBS participants are well characterized and followed up for over three years, allowing the investigation of the metabolomic effects alongside a wide range of clinical phenotypes. However, BBS participants are predominantly white and of European ancestry and therefore the data are limited in their generalisability to other populations beyond the UK. Many of the baseline (pre-surgery) samples do not have a corresponding post-surgery sample from the same participant, which reduces power to detect metabolomic change associated with bariatric surgery. However, the unpaired baseline samples can still contribute power to statistical models. The NMR platform used measured of biomarkers with strong clinical relevance including many lipid subtypes. Combining these data with untargeted mass spectrometry-based data in the same sample^6^ delivers a wide coverage of the metabolome. A key strength of the NMR data is the absolute concentration units which are physiologically meaningful and allow comparison across sample sets and studies.

## Conclusion

To summarize, we have provided an overview of NMR data generated in BBS, a clinical trial comparing three types of bariatric surgery. QC procedures and some data validation analyses have been described. Future users of the data may wish to apply additional QC and validation procedures specific to their analytical requirements. The filtered dataset consists of 1410 samples with targeted NMR metabolomics data alongside comprehensive clinical characteristics. The dataset provides a useful tool for exploring the metabolomic effects of bariatric surgery and its concurrent weight loss.

## Data Availability

At the time these data were generated, participants were not asked for their permission to share data beyond the immediate project team. However, anonymised individual patient data from the By-Band-Sleeve trial will be made available upon request to the chief investigator (Jane Blazeby) for secondary research, conditional on assurance from the secondary researcher that the proposed use of the data is compliant with the Medical Research Council Policy on Data Sharing regarding scientific quality, ethical requirements, and value for money, and is compliant with the National Institute for Health and Care Research policy on data sharing. A minimum requirement with respect to scientific quality will be a publicly available prespecified protocol describing the purpose, methods, and analysis of the secondary research (e.g., a protocol for a Cochrane systematic review), approved by a UK Research Ethics Committee or other similar, approved ethics review body. Participant identifiers will not be passed on to any third party.

https://osf.io/uwehr/

https://github.com/lauracorbin/bbs_nmr_datanote

## Data and code availability

### Dataset

One dataset has been generated as part of this work:

### Extended data

Open Science Framework: Supplementary information supporting this submission can be found on the Open Science Framework “Quantitative biomarker profiling of serum samples in the By-Band-Sleeve trial” project page (DOI: 10.17605/OSF.IO/UWEHR).

This project contains the following extended data:

- ‘SF1-224375 CorbinCT-26-Jul-2023-Results-overview.pdf’ is the results overview provided by Nightingale Health to accompany the data provided.
- ‘SF2-224375 CorbinCT-28-Jul-2023-Quality-report.pdf’ is the quality report provided by Nightingale Health to accompany the data provided.
- ‘SF3-figure_S1.pdf’ contains Bland-Altman plots of the duplicated sample pairs. Only metabolites with units in mmol/L are plotted.
- ‘SF4-table_S1.xlsx’ - student’s t test results per metabolite for the suspected outlier samples versus the rest of the samples.
- ‘SF5-metaboprep_summary_report.html’ – is the *metaboprep* summary report for the final QC’d dataset comprising 1410 samples.
- ‘SF6-figure_S2.pdf’ – Bland-Altman plots of clinical and NMR measures after removal of samples with ‘low protein’ flag.
- ‘SF7-figure_S3.pdf’ - contains Bland-Altman plots for each metabolite comparing the 2022 pilot analysis to the 2023 analysis, for a subset of 250 samples collected at site B.

Data are available under the terms of the Creative Commons Attribution 4.0 International license (CC-BY 4.0).

### Code

Code used to derive and validate the dataset described is openly available in GitHub at: https://github.com/lauracorbin/bbs_nmr_datanote (DOI: 10.5281/zenodo.17198828).

## Author contributions

Conceptualisation: LJC, NJT; Data curation: MLS, LJC, EAG; Formal analysis: MLS, LJC, DAH; Funding acquisition: NJT, JMB, CAR; Investigation: JMB, CAR (leads of BBS study); Resources: SF, AG, SMR, JMB, CAR, GM, EAG; Supervision: LJC, NJT; Validation: MLS, DAH; Visualisation: MLS, DAH; Writing – original draft preparation: MLS; Writing – reviewing & editing: MLS, LJC, LJG, DAH, NJT, CAR, JMB.

## Acknowledgements

The By-Band-Sleeve trial is led by JM Blazeby (Chief Investigator) and CA Rogers (lead methodologist). We acknowledge the support of all other members of the NIHR By-Band-Sleeve Management Group: Robert C Andrews, John Bessant, James P Byrne, Nicholas Carter, Caroline Clay, Jenny L Donovan, Eleanor Gidman, Graziella Mazza, Mary O’Kane, Barnaby C Reeves, Nicki Salter, Janice L Tompson, Richard Welbourn and Sarah Wordsworth. We also thank all the By-Band-Sleeve contributors including the investigators, research dieticians and nurses, the independent trial steering committee and data monitoring and safety committee. We are grateful to all the patients who participated in this trial.

We thank the technical team at the Bristol Bioresource Laboratories at the University of Bristol for sample management once samples had been processed.

We thank the technical team at Nightingale Health for their advice regarding the data validation results.

